# Nanopore sequencing-based episignature detection

**DOI:** 10.1101/2024.04.19.24305959

**Authors:** Mathilde Geysens, Benjamin Huremagic, Erika Souche, Jeroen Breckpot, Koenraad Devriendt, Hilde Peeters, Griet Van Buggenhout, Hilde Van Esch, Kris Van Den Bogaert, Joris Robert Vermeesch

## Abstract

**Background:** A subset of developmental disorders (DD) is characterized by disease-specific genome-wide methylation changes. These episignatures inform about underlying pathogenic mechanisms and can be used to assess the pathogenicity of genomic variants as well as confirm clinical diagnoses. Currently, episignature detection requires the use of indirect methylation profiling microarrays. We hypothesized that long-read whole genome sequencing would not only enable the detection of single nucleotide variants and structural variants but also episignatures.

**Methods:** Genome-wide nanopore sequencing was performed in forty controls and twenty patients with confirmed or suspected episignature-associated DD, representing thirteen distinct diseases. Following variant and methylome calling, hierarchical clustering and dimensional reduction were used to determine the compatibility with microarray-based episignatures. Subsequently, we developed a support vector machine for each DD.

**Results:** Nanopore sequencing based methylome patterns were concordant with microarray-based episignatures. Our classifier identified episignatures in 17/20 disease samples and none of the controls. The remaining three patient samples were classified as controls by both our classifier and a commercial microarray assay. In addition, we identified all underlying pathogenic single nucleotide and structural variants and showed haplotype-aware skewed X-inactivation evaluation directs clinical interpretation.

**Conclusion:** This proof-of-concept study demonstrates nanopore sequencing enables concurrent haplotyped genomic and epigenomic analyses.

## INTRODUCTION

The epigenome plays a central role in regulating differential gene expression. Epigenetic variation is therefore involved in several adaptative but also pathological processes. Repeat expansion-associated modifications of promotor methylation have long been recognized as a cause of developmental disorders [1], [2]. Imprinting defects are also known to disturb development, growth, and metabolism [3]. More recently, methylome studies in patients with unexplained DD revealed rare epigenetic changes in 23% of patients with a 2.8 fold excess of *de novo* epivariants [4], suggesting that these may be involved in their etiology. Yet, apart from a handful of targeted methylation assays for well-known imprinted regions, epigenomic analyses are currently not routinely performed in the diagnostic work-up of patients with DD.

Besides localized epigenetic variants, genome-wide methylation studies in DD have recently revealed multi-loci, disorder-specific methylation disturbances, called episignatures. Episignatures have initially been identified in disorders caused by known chromatin and/or methylation regulatory gene disruptions [5]. An increasing number of disorders are characterized by episignatures and recognizable methylation pattern changes have now been detected in over sixty diseases [6]. For some, such as certain microdeletion syndromes [7], the mechanism causing reproducible genome wide methylation changes remains unclear. Mapping the methylome in DD will therefore improve our understanding of pathogenic mechanisms and might leverage new insights in the origins of phenotypic variability. In addition, episignatures have a high diagnostic value in assessing the pathogenicity of genomic variants of unknown significance (VUS) and in confirming clinical diagnoses in patients for which the underlying genomic variant could not be identified [8].

The current method used to identify episignatures is based on bisulfite conversion of methylated cytosines followed by methylation analysis using Illumina Epic/Infinium methylation microarrays [6] and commercialized as the EpiSign assay. A clinical implementation study of this method reported the detection of an episignature in 11% of patients with DD without conclusive genetic finding and in 35% of patients with a VUS in a gene for which an episignature is described [9], demonstrating its diagnostic value. One drawback of this assay is that it represents an extra step in the diagnostic odyssey of patients which comes with additional diagnostic delays and increased costs.

With the advent of long-read sequencing technologies, detection of single nucleotide variants (SNVs), structural variants (SVs), and base modifications in a single assay becomes reality [10]. Both prevailing long-read methodologies, nanopore (Oxford Nanopore Technologies) and single molecule real-time sequencing (PacBio), enable methylation detection from the native DNA strands without amplification, bisulfite or enzymatic conversion biases [11]. Moreover, the generated long reads enable haplotyping [12], allowing the phasing of genetic and epigenetic variation. Recent studies showed the ability of nanopore sequencing to detect X-chromosome inactivation (XCI) [13] as well as methylation disturbances at imprinted loci [14] and short tandem repeats causing DD [15].

Considering the potential of nanopore sequencing to detect localized methylation levels and disease states, we reasoned it should be possible to map methylome-wide disturbances in chromatinopathies. Hence, we explored the potential to concurrently detect episignatures and underlying pathogenic variants. We performed long-read whole genome nanopore sequencing (lrWGS) of twenty patients representing thirteen different rare DD with known episignatures, as well as forty controls. We report non-inferiority of nanopore sequencing for detecting episignatures and demonstrate concurrent detection of underlying SNVs or SVs. We also confirm the detection of imprinting as well as haplotype-aware skewed XCI. In addition, our results illustrate some limitations of current episignatures. Overall, our study underscores the diagnostic value of lrWGS which might ease rare diseases diagnosis by offering simultaneous genome and epigenome analysis.

## MATERIAL AND METHODS

### Patients’ inclusion and sequencing

Twenty patients with a confirmed or a suspected DD for which an episignature is described (Table 1, Suppl. Table 1), as well as forty healthy controls, were included. For the first phase of our study, we included eleven patients representing four diseases as well as five controls. For the second phase, nine additional patients with different diseases were included to increase the amount of evaluated episignatures as well as thirty-five other controls. Two of the patients were selected based on the presence of a VUS and not a (likely) pathogenic variant, as evaluation and illustration of one of the major applications of episignature detection. The genomic variants of all patients had been identified by chromosomal microarray, targeted Sanger sequencing or trio-based exome sequencing (WES).

**Table 1.**
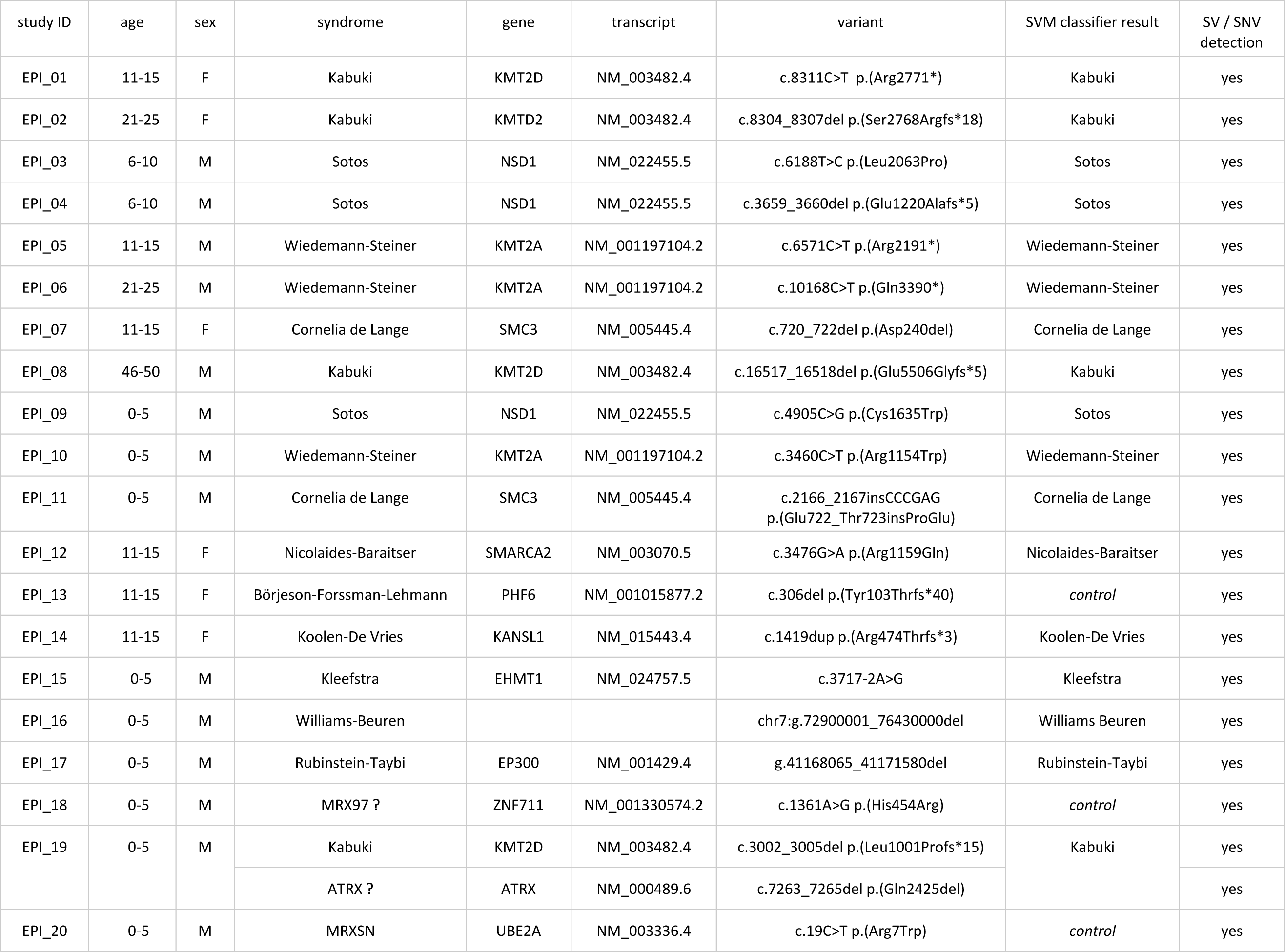
Concurrent detection of episignatures (SVM based) and underlying genomic variants. The seven first columns of the table describe patient characteristics and (likely) pathogenic genomic variants or VUS (marked by a “?”) detected by standard of care methods. The next column indicates the result of the SVM based classifier for each sample. Patients for which no episignature is identified by the classifiers are classified as control (italic). The last column specifies if SNV calling (Clair3) or SV calling (Sniffles2 and QDNAseq) allowed the identification of the genomic variant by nanopore sequencing.

Fresh or frozen (−80°C) blood was used for HMW DNA extraction (Promega Wizard®, Monarch®). For four patients and two controls, DNA extracted with the automated Revvity – Chemagic® workflow and stored at −20°C was retrieved (Suppl. Table 2). Libraries were prepared from 3µg of DNA using the Oxford Nanopore Technologies (ONT) Ligation Sequencing Kit (SQK-LSK110). Each sample was loaded on a R.9.4.1 Flow Cell and reloaded once after 24-48h, for a total sequencing period of 72-96h on a PromethION®. From HMW DNA extracted from fresh blood, we obtained a median output of 95.6 GB and a median N50 of 40.4 kb. Using frozen blood, the median output was 116.25 GB, and the median N50 31.9 kb. Using stored non-HMW DNA, the median output and N50 were 113.7 GB and 17 kb, respectively (Suppl. Table 2).

For patient EPI_13, additional urine and buccal swab samples were collected. DNA extraction of these tissues was performed using the Qiagen® and MagCore® chemistries, respectively. Using the SQK-LSK110 chemistry and r.9.4.1 Flow Cells we generated an output of 100 GB for the urine (N50 = 9.79 kb) and 146.7 GB for the buccal swab (N50 = 7.25 kb).

### (Modified) basecalling, mapping, SVs and SNVs calling

To extract methylation information from raw ONT data, we performed modified basecalling with Dorado (v0.3.0) using the dna_r9.4.1_e8_sup@v3.3_5mCG model[17]. After basecalling, reads were aligned to hg38 with minimap2 (v2.24) [18]. Subsequently, bedMethyl files, used in downstream analysis, were generated from aligned BAM files using modkit (v0.1.13 with pileup --preset traditional --only-tabs) [19].

Ultimately, starting from aligned BAMs, SVs were called using Sniffles2 (v2.0.2) [16] and QDNAseq (v1.3.8) [17]. Large copy number variants (>50 kb) are known to be difficult to detect with current nanopore SV callers such as Sniffles2 [18], and read-depth based softwares, such as QDNAseq, can therefore be used to improve the sensitivity for these variants [19]. SNVs were called with Clair3 (v1.0.4)[20], which was also used to generate haplotagged BAM files. Methylation was visualized with Methylartist [21] and Integrative Genomics Viewer (IGV) [22].

### Detection of differentially methylated regions: imprinting and X-inactivation

Imprinting was assessed by quantifying and visualizing methylation of both haplotypes at six loci located in three imprinted regions associated with DD (11p15.5, 14q32, and 15q11-q13). The following CpGs were assessed: 242 CpGs at the *H19* (chr11:1997509-2003349, hg38), 194 CpG at the *KCNQ1* (chr11:2698155-2701028, hg38), 188 CpG at the *MEG3* (chr14:100824185-100827640, hg38), 45 CpG at the *MEG8* (chr14:100904325-100905081, hg38), 116 CpG at the *SNRPN* (chr15:24954564-24956828, hg38) and 52 CpG at the *MAGEL2* (chr15:23647178-23648424, hg38) imprinted loci.

To evaluate XCI, we assessed methylation at two validated loci, also recently targeted by CRISPR-Cas9 nanopore sequencing for this purpose: 115 CpGs at the *AR* gene loci (chrX:67543761-67546170, hg38) and 57 CpGs at the *RP2* gene loci (chrX:46836539-46837273, hg38) [23]. In addition, 99 CpGs in the promotor region of *PHF6* were evaluated (chrX:134372110-134374891, hg38).

Differential methylation at these loci was quantified and visualized in the six females of our cohort. Loci with <10x total coverage or <5x coverage for one of the haplotypes were excluded from subsequent analyses (3/18 loci for both imprinting and XCI analyses).

For the mother of EPI_19, XCI was evaluated by the standard of care method, *i.e.* comparing the relative amount of PCR amplification products of both *AR* gene CAG-repeat haplotypes after methylation-sensitive restriction enzyme digestion.

### Distinction of episignatures by dimensional reduction and clustering

Thirty-four disease-specific episignatures have been identified through microarrays and published by E. Aref-Eshghi *et al.* [4]. The most significant microarray probes for each disease and methylation levels at these loci have been made available. We extracted the DNA methylation beta values from this published dataset at all episignatures probes’ loci for the thirty-four diseases and the included control. This extraction provided, for every episignature locus, one array-based example of the episignature in presence of the disorder and thirty-four examples of methylation levels at these loci in the absence of the disorder. Using pyliftover [24], all the probes (loci) encoding episignatures were lifted from hg19 to hg38. During the liftover, only a single CpG locus in the SBBYSS episignature was lost. Methylation levels were extracted from the nanopore sequencing data at the episignature loci using hg38 coordinates. During this process, the percentage of methylated reads was extracted from bedMethyl files. Using python (v3.9.15), three approaches were used to determine the similarity between array- and nanopore-based episignatures: UMAP (umap-learn_v0.5.4, n_neighbors=2) and t-SNE (sklearn_v1.2.2, n_components=2, perplexity=2) for dimensional reduction, and hierarchical clustering (scipy_v1.9.3, seaborn_v0.12.2).

### Development of a SVM classifier

We adopted the One-vs-All approach and trained 34 individual Support Vector Machines (SVMs) using disorder episignature loci DNA methylation median beta values [4]. Every SVM (sklearn v1.2.2) was trained to predict the presence of a specific disorder episignature considering all the other cases as controls. These SVMs were used to classify the nanopore-based methylome profiles. To predict the sample’s class, we ran all the classifiers and assigned the sample to the class with the highest confidence score. Confidence scores represent the decision function indicating the signed distance of a sample from the separating hyperplane. If none of the classifiers returned a positive confidence score or all the confidence scores were <0.30, the sample was classified as control.

While this approach guaranteed one and thirty-four examples of methylation values in the presence and in the absence of the disorder, respectively, it also introduced an unbalance between the positive (disorder presence) and negative (disorder absence) classes that could affect the SVM training. To overcome this issue, we introduced a parameter adjusting the weight of the classes during the training of SVMs. The weight for the positive class training sample was set to ten, and the weight for the negative class training samples to one.

## RESULTS

### Imprinting detection

To verify the haplotype-aware methylation detection, six loci localized in three imprinted regions (11p15.5, 14q32, and 15q11-q13) were investigated in six patients (Figure 1, Suppl. Figure 1A). Visualizing both haplotypes’ methylation, we observe consistent methylation of most CpGs of one allele in contrast to the absence of methylation of most CpGs of the other haplotype at all loci in all patients. The *H19* and *SNRPN* loci in EPI_12, and *H19* locus in EPI_01, could not be assessed given the low coverage. Further quantification of methylation at these loci shows mean percentages of methylation between 89.3% and 96.3% for the methylated haplotypes and 4.1% to 16.5% for the unmethylated alleles (Suppl. Figure 1A, Suppl. Table 3A). These results are concordant with expectations of mono-allelic expression at imprinted loci and illustrate direct allelic methylation measurement as comparison to standard technologies where disturbances of one allele’s methylation are inferred from total methylation levels.

**Figure 1.**
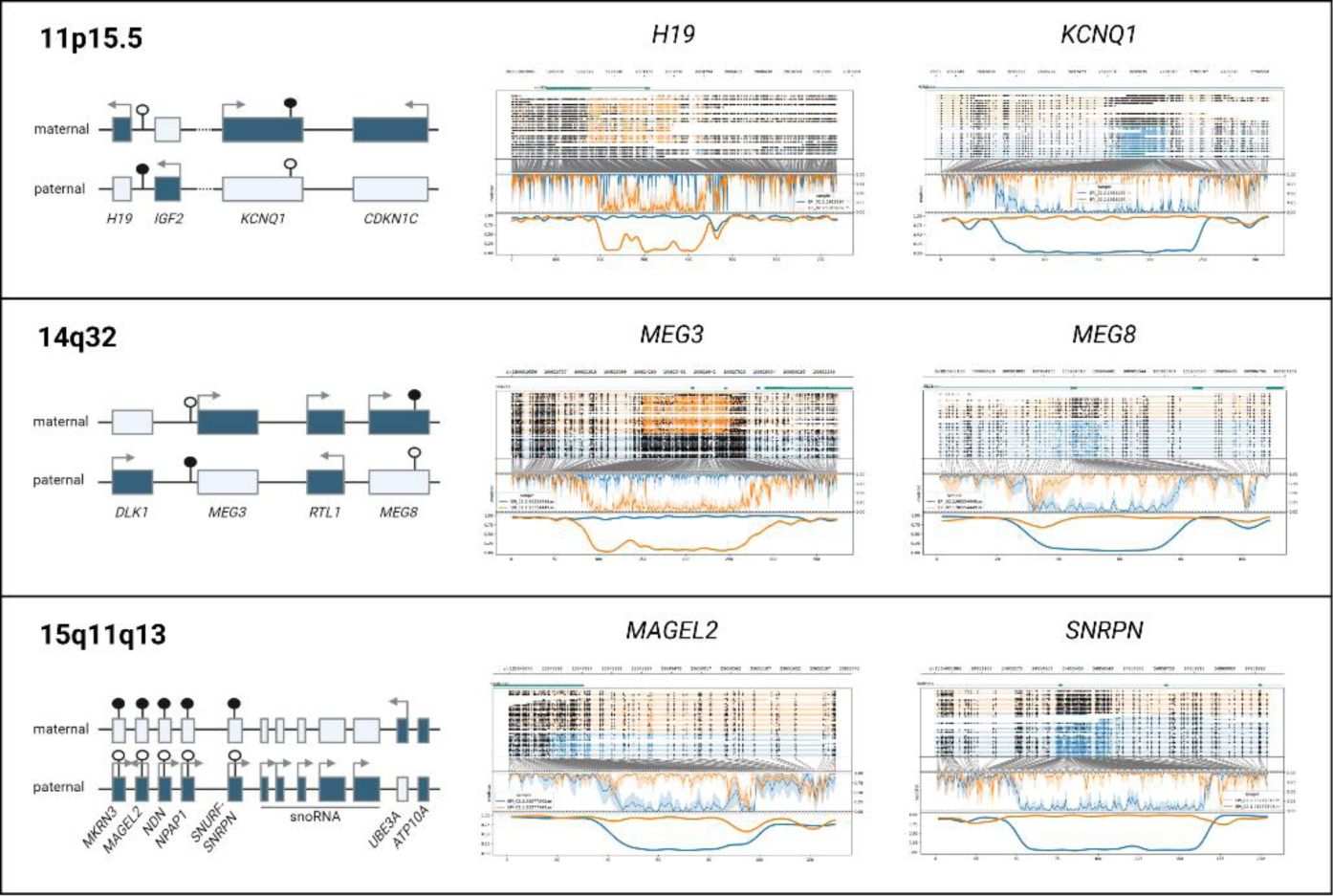
Detection of differential methylation at imprinted loci. Haplotype aware methylation visualization of EPI_02 at 6 imprinted loci (middle and right part). The maternal allele is represented in orange and paternal allele in blue. The upper part of each plot represents the reads, with methylated CpGs visualized as black circles. In the lower part of each plot, after translation from genome to CpG-only coordinate space, raw log-likelihood ratios are plotted above a smoothened graph of methylation fraction. The left part illustrates gene localization and expression at these 3 imprinted loci. Active genes are represented as full boxes with arrows, whereas untranslated genes are represented by empty boxes. Full circles represent methylated CpG islands whereas empty circles represent unmethylated CpG islands.

### Episignature distinction using microarray data as reference

To evaluate the potential of nanopore sequencing-based episignature detection, seven patients were initially sequenced: two with Kabuki, Sotos, and Wiedemann-Steiner syndrome, and one with Cornelia de Lange syndrome. Two-dimensional reduction analyses of episignatures (Figure 2A, Suppl. Figure 2), comparing our samples to the microarray-based disease reference as well as five healthy controls, co-located nanopore methylation values close to the reference microarray data for each disease sample at their disease-specific loci.

**Figure 2.**
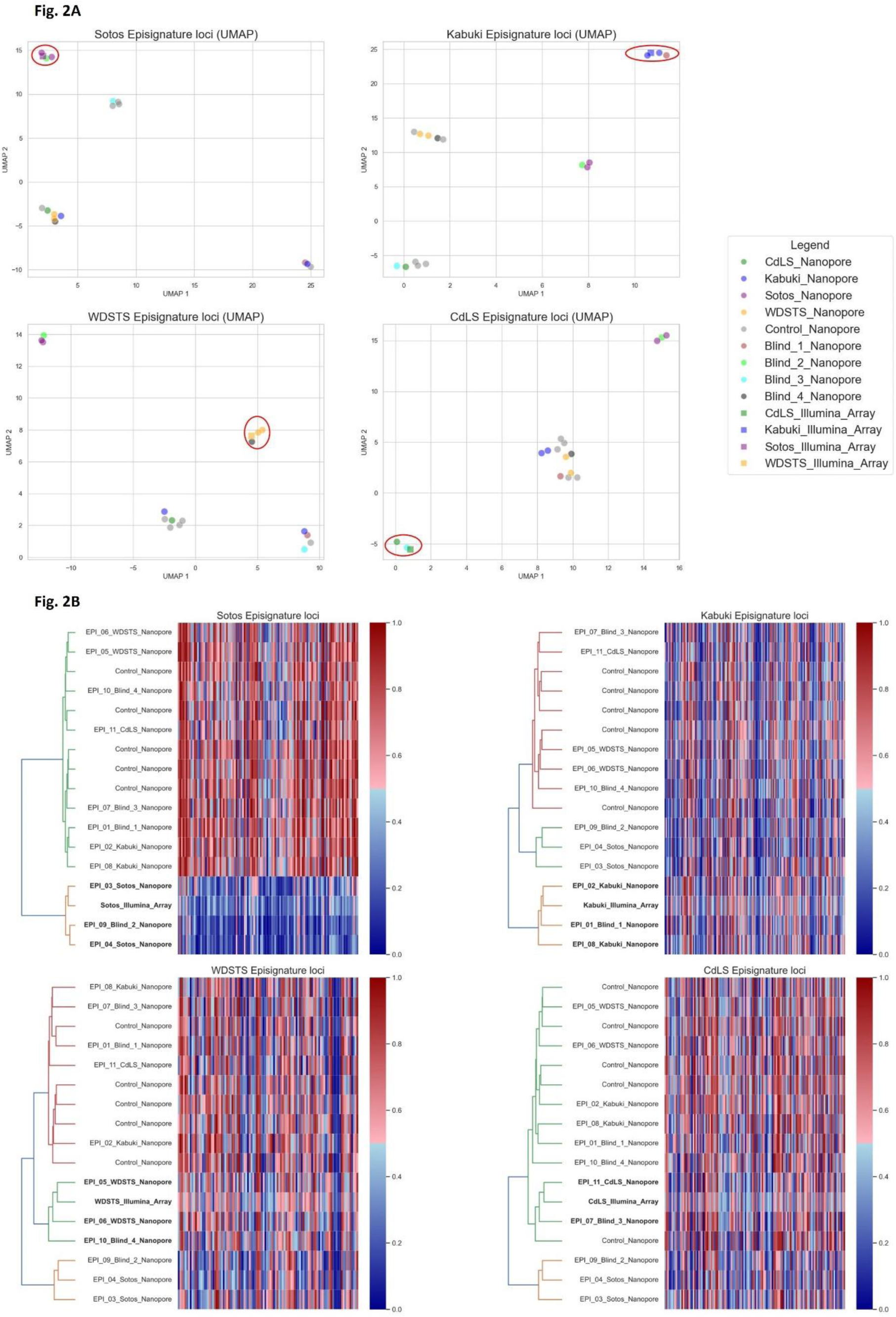
Nanopore methylation analysis allow to distinguish disease samples from controls and other diseases samples. UMAP scatter plots (Fig. 2A), hierarchical clustering (Fig. 2B, left part) and heatmaps (Fig. 2B, right part) based on methylation levels at Sotos, Kabuki, Wiedemann-Steiner (WDSTS) and Cornelia de Lange (CdLS) syndromes episignature loci. Nanopore sequencing data of five healthy controls and eleven (seven known and four blinded) patients with one of the four disorders are represented, along with the array-based reference for the respective episignature. Patients sequenced with nanopore cluster in close proximity to their respective array-based episignature reference. Blinded samples can be assigned the correct diagnosis.

To further confirm the hypothesis that nanopore-based methylome analysis can be used to recognize episignatures, we sequenced an additional four blinded cases (one of each disease). Hierarchical clustering (Figure 2B) of methylation levels at the disease’s episignature loci combined with the microarray methylome profiles used as a reference, clustered all blinded samples with the disease reference and other samples with the same disorder. Similarly, the blinded samples clustered with samples with the same disease in the UMAP analysis (Figure 2A). Methylation levels at other disease-specific loci are discriminative for some diseases (*e.g.* CdLS and WDSTS loci for Sotos), but not for all (*e.g.* Kabuki loci for Sotos), as was shown in the original publications [25]. Moreover, the heatmaps show how strong the methylation differences are in some diseases (*e.g.* Sotos), but subtle in others. Nonetheless, even in those diseases with small methylation variation, hierarchical clustering enables to recognize each samples’ disease.

### SVM based episignature detection

Our primary results showed, through different approaches, a high similarity between the microarray reference and nanopore episignatures. However, methods like UMAP require interpretation upon visual representation, and in the case of hierarchical clustering, determining the cutoffs or number of clusters is not trivial. For this reason, we developed an automated and generic approach to screen samples for 34 shared episignatures [5] using SVM classifiers. Our classifiers were tested on the eleven patients assessed in our first analyses and nine additional samples, representing thirteen diseases, as well as forty controls. In 17/20 patients with a (suspected) DD, the classifiers recognized an episignature and assigned the sample to the right disease (Table 1). All healthy individuals were classified as controls.

For patient EPI_19, both a maternally inherited, hemizygous c.7263_7265del p.(Gln2425del) VUS in *ATRX* and a pathogenic but mosaic c.3002_3005del p.(Leu1001Profs*15) variant in *KMT2D* (present in 10-21% of cells according to WES and Sanger sequencing) were identified. The detection of the mosaic *KMT2D* variant was driven by clinical suspicion, but an additional ATRX syndrome could not be excluded solely based on the patient’s phenotype. The SVMs classified this patient as Kabuki syndrome, the syndrome caused by pathogenic variants in *KMT2D,* but not as ATRX syndrome. Interestingly, the confidence score value returned by the SVM was lower compared to other Kabuki patients: 0.64 in EPI_19 in contrast to 1.94, 1.78, and 2.30 for the three other Kabuki samples (Suppl. Table 4), likely as consequence of the mosaic status of the *KMT2D* variant. Similarly, EpiSign also identified the Kabuki and not the ATRX signature. In addition, XCI testing was performed in the mother and showed a 41/59 inactivation ratio. Hence, there is no skewed XCI in maternal blood. Moreover, segregation analysis showed the presence of the *ATRX* c.7263_7265del p.(Gln2425del) variant in a healthy brother of the patient, further supporting the *ATRX* variant to be benign. Together with the result of our classifier, those results indicate the *KMT2D* c.3002_3005del p.(Leu1001Profs*15) variant to be pathogenic, but not the *ATRX* c.7263_7265del p.(Gln2425del) variant.

In three samples (EPI_13, EPI_18 and EPI20), the classifier did not recognize any of the reference episignatures (Table 1). For those cases, clinical testing using Episign also returned negative.

### Supporting evidence from phased X-inactivation analysis

Patient EPI_13 had a diagnosis of Börjeson-Forssman-Lehmann syndrome (BFLS), after identification of a *de novo* pathogenic c.306del p.(Tyr103Thrfs*40) variant in *PHF6*. BFLS is an X-linked disorder with variable penetrance and expression in females. Functional mosaicism, a phenomenon where the allele on which the pathogenic variant is located is active in some but not all cells due to XCI, can explain, at least in part, this variation [26]. Hence, we hypothesize that the female patient with BFLS (EPI_13) might express the allele with the pathogenic variant in relevant tissues but have skewed XCI in blood with predominant inactivation of the abnormal allele. This could explain the absence of episignature in blood. Methylation analysis of the two *AR*, *RP2*, and *PHF6* promotor haplotypes on the patient’s blood sample showed divergent methylation levels between both haplotypes: 75%:2% at the *AR* loci, 80%:5% at the *RP2* loci and 79%:10% at the *PHF6* loci (coverage of 37x for the three loci). These results point to skewing of XCI. In comparison, 26% to 68% of the reads of each of the two haplotypes are methylated at these loci in the five other females of the cohort (Figure 3, Suppl. Table 3B). Evaluation of XCI of the patient’s urine and buccal swab samples also revealed an imbalanced inactivation, the percentages of methylation of both alleles being measured at 76%:19% (*AR*) and 75%:12% (*RP2*) for the urine sample and 71%:3% (*AR*) and 83%:3% (*RP2)* for the buccal swab sample (Suppl. Figure 1B).

**Figure 3.**
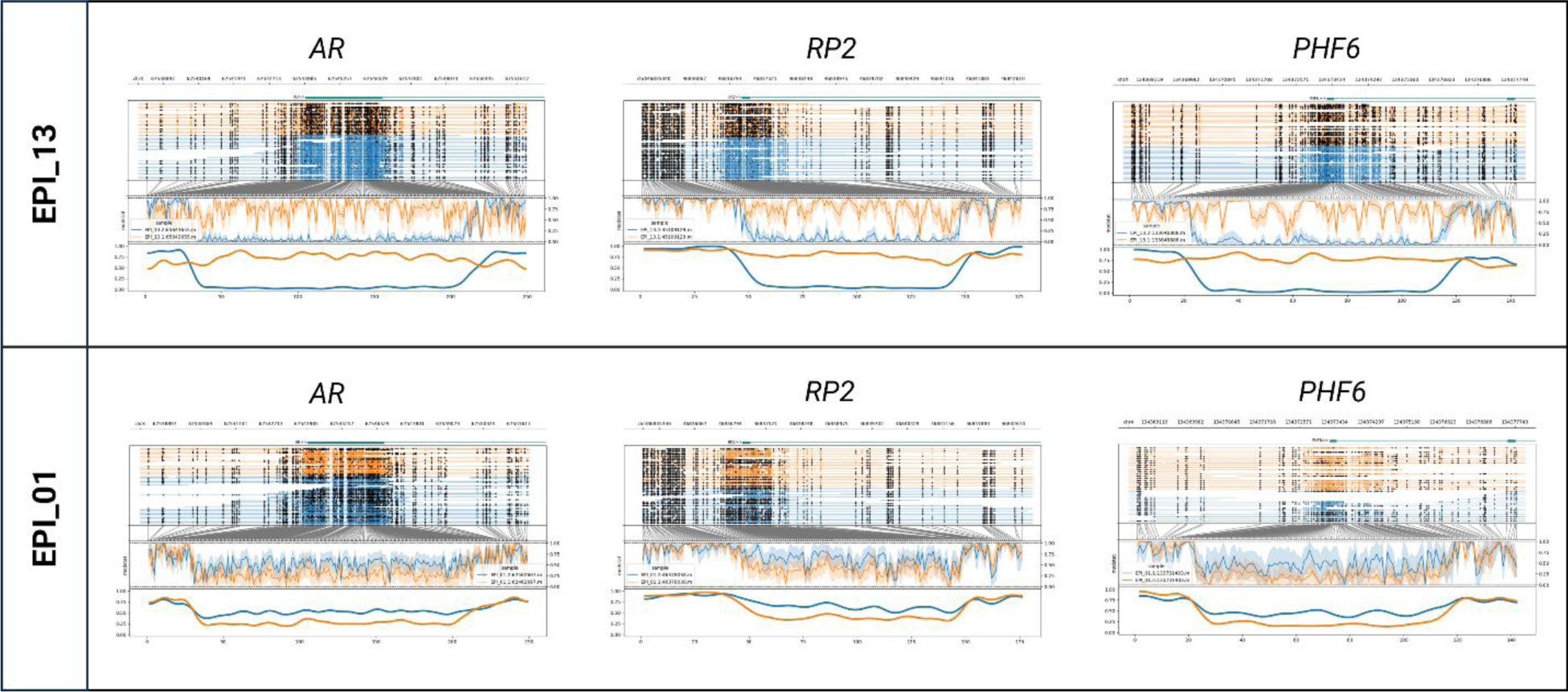
Evaluation of X-inactivation. Phased nanopore sequencing methylation levels at AR and RP2 loci, as well as at the promotor of the PHF6 gene, in patient EPI_13 (top) and EPI_01 (bottom). Orange and blue represent the two different haplotypes of each sample. In the upper part of each plot, the genome coordinates, genes and reads are illustrated. On each read, modified bases are represented by full, dark circles, unmodified bases by open circles. In the lower part of the plot, after translation from genome to CpG-only coordinate space, raw log-likelihood ratios are plotted above a smoothened graph of methylation fraction. About half reads of each allele are methylated at the three loci in patient EPI_01 whereas in patient EPI_13 one allele (orange) is methylated in all reads in contrast to the other one which is unmethylated.

To further substantiate our hypothesis, we confirmed EPI_13’s *PHF6* c.306del p.(Tyr103Thrfs*40) pathogenic variant to be localized on the hypermethylated inactive allele by haplotyping. Phasing is enabled by the long reads spanning the 5’UTR differentially methylated region to the pathogenic variant. Taking further advantage of this property, we could determine that the *de novo PHF6* pathogenic variant occurred on the maternal allele. Available trio WES data showed an informative maternally inherited SNV 521 base pairs downstream of the pathogenic variant and nanopore long reads co-localized these two variants to be in *cis* (Suppl. Fig. 3). The *RP2* locus did not contain any informative SNV. The *AR* differentially methylated region, however, also contained an informative SNV which confirms the inactivation of the mutated maternal allele. These results support the hypothesis that this variant is pathogenic but, due to skewed X-inactivation, is not causing an epigenetic fingerprint in white blood cells.

### Concurrent epigenetic and genetic diagnosis

Given that long-read sequencing offers genome in parallel with methylome sequencing, we wanted to evaluate and confirm the potential of long-read sequencing to enable concurrent episignature, SV, and SNV detection. Hence, we assessed whether the (likely) pathogenic variants (or VUS for EPI_18 and EPI_19) could be detected in the nanopore sequencing data. Eighteen of nineteen SNVs were identified using the standard bioinformatic analysis (see M&M). The mosaic *KMT2D* variant in EPI_19 was only present in 17% (5/29) of the nanopore reads and could not be detected. However, the variant allele frequency in previous short-read WES was also too low (22%, 38/174 reads) to be detected after default variant allele frequency-based filtering. Both short- and long-read methods did allow the *KMT2D* variant detection after manual curation driven by clinical suspicion. Two patients were heterozygous for a pathogenic SV and both were detected: the multi-exonic deletion in the *EP300* gene in EPI_17 and the 7q11.23 microdeletion in EPI_16. Additionally, the long-read sequencing data allowed mapping the breakpoints of the *EP300* deletion (NM_001429: g.41168065_41171580del, hg38).

## DISCUSSION

Episignatures are diagnostically important. Their presence provides evidence for the pathogenicity of VUS or can act as biomarker for a disease when the underlying genomic variant cannot be identified [6], [8]. This proof-of-concept study illustrates the potential of lrWGS for episignature detection. Despite known technological differences, we show that microarray reference data can be used for several diseases. Moreover, nanopore sequencing enables to concurrently identify the episignature inducing SNVs and SVs. In addition, we show that phased methylation maps inform about imprinting and skewed XCI. Altogether, long-read sequencing haplotype-aware mapping of genetic and methylation variation provides a comprehensive genome analysis, combining several analyses that currently require multiple technologies.

While this study opens the door for concurrent genomic and episignature assessment, it also illustrates limitations of current episignatures. For two of the genes of which the episignature could not be detected, *UBE2A* and *PHF6,* the reference microarray data we used are based on a limited sample set [5] without test nor validation cohort, and might therefore be imprecise. Both genes, as well as *ZNF711*, are also listed among the genes for which the commercial microarray assay is reported to have a lower sensitivity [27]. Moreover, in the last years, sub-episignatures have been described for different types of variants in the same gene [28],[29]. While not yet been reported for *UBE2A* and *ZNF711*, such sub-episignatures could confound the classification of EPI 18 and EPI_20, which both harbor missense variants. Segregation, maternal XCI analyses, and functional analyses performed in a previous study have confirmed the pathogenicity of the c.19C>T p.(Arg7Trp) variant in *UBE2A* identified in *EPI_20* [30]. Segregation and functional analyses could not be performed for the c.1223A>G p.(His408Arg) VUS in *ZNF711* identified in EPI_18. In this case, the absence of the disease-associated episignature might therefore indicate the variant to be benign. However, one must carefully draw conclusions based on the absence of episignatures as a sub-episignature cannot be excluded. An evaluation of published episignatures also recently showed their high specificity but variable sensitivity [31].

Another constraint highlighted by this study is XCI. For the *PHF6* c.306del p.(Tyr103Thrfs*40) variant (EPI_13), we hypothesized that skewed XCI in white blood cells might lead to predominant expression of the normal allele, resulting in a normal methylation pattern. Skewed XCI has already been reported in other females with BFLS [32]. Using haplotype-aware nanopore methylation mapping, we confirmed the unbalanced skewing in this patient and subsequently localized the variant on the inactivated allele. As the patient presents high phenotypic similarities with BFLS, we hypothesized that other tissues probably lack XCI skewing and might present the episignature. However, both urine and buccal swab samples also showed predominant inactivation of the affected allele. Other tissues could be investigated to substantiate our hypothesis further, but this requires more invasive procedures and might require tissue-specific epigenomic maps, which are currently lacking. Concurrent XCI and episignature evaluation in blood of carrier mothers of males with a X-linked disease episignature could, therefore, provide an indirect alternative to validate this hypothesis in the future.

An important potential pitfall in the diagnostic process of DD is mosaicism. Post-zygotic *de novo* pathogenic variants can lead to mosaic variants which often escape detection by the default variant calling algorithms. Interestingly, our classifier did detect the episignature in a mosaic heterozygous patient (EPI_19), albeit with a lower confidence score, reflecting the low-grade mosaicism for the pathogenic variant. Mosaicism detection can be improved by including mosaic samples in the training set and lowering the positive confidence score threshold [33]. Episignature evaluation was important in this boy as it contributed to classifying the X-linked *ATRX* variant as benign and reassuring the parents of low recurrence risk for future pregnancies. The *ATRX* episignature has indeed been reported as having a high sensitivity [31], and this conclusion was corroborated by segregation and maternal XCI analyses. We expect that episignature detection can also potentially direct the clinical diagnostic laboratory to search for mosaic variants in specific genes.

The correlation between microarray- and nanopore-based methylomes has been estimated at around 0.85 [34]. Here we show that despite these inter-assay biases and the subtle methylation changes that characterize the episignatures of some disorders, publicly available microarray-based methylation ratio profiles and/or positions can be used for nanopore sequencing episignature detection for several disorders. Still, it is likely that comparing nanopore methylation to microarray reference data reduces the sensitivity. Microarray references also have the disadvantage of being based on a restricted amount of CpGs, varying with the product’s version and each sample’s hybridization [35]. We envision that large-scale nanopore sequencing of patients and controls in the future will leverage high-resolution whole methylomes, enabling refinement of existing episignatures, potentially improving their sensitivity and specificity, and likely leading to the discovery of new episignatures.

Important drawbacks of LRS platforms have long been their lower accuracy and high costs. However, the costs of nanopore sequencing have been dropping and currently (for 30x, genome-wide LRS) slightly exceed the costs of clinical episignature testing by microarrays in a large-scale setting. With the improving accuracy of LRS chemistries and analytical tools, the cost-effectiveness balance might change as concurrent genetic and epigenetic testing would enable to replace the combination of WES and methylation-sensitive microarray analyses. The other side of the coin of all the analyses enabled by LRS remains the high amount of resources that are needed to store, process and analyze the data.

To conclude, lrWGS enables concurrent phased genome and methylome variant detection, opening the door to large-scale studies about genomic and epigenomic variation and their interactions. While this study has used nanopore sequencing, other sequencing platforms combining methylation and base calling will probably produce similar results. XCI and imprinting detection have already been demonstrated with single-molecule real-time sequencing too [36], [37]. With its improving SNV calling accuracy [10] and more comprehensive SV detection, it seems likely that long-read sequencing will be implemented as the first-tier test for the diagnosis of DD in the future. Beyond known and potential new episignatures, we envision that shedding light on secondary epivariation will improve our understanding of genomic, especially non-coding, variants, and give us new insights into molecular mechanisms underlying diseases. In addition, an excess of *de novo* primary epivariants has been identified in patients with unexplained DD [4], and the presence of such epivariants has been associated with outlier gene expression [1]. Further exploration of the whole epigenome might, therefore, help us solve another part of the remaining unexplained DD.

## Data Availability

Data produced are partially available as supplementary with the manuscript and on GitHub.

https://github.com/JorisVermeeschLab/NSBEpi

## DATA AVAILABILITY

Methylation values at used genomic positions are shared in supplementary table 5. The used data and code are available on GitHub: JorisVermeeschLab/NSBEpi: Nanopore sequencing-based episignature detection (github.com). Bedmethyl files are available upon request.

## ACKNOWLEDGEMENTS

The authors thank the patients and their families who participated in this research. We thank Jonas Demeulemeester for sharing his expertise about nanopore sequencing methylation calling at the start of the project and also thank Anne Bassett and Donna McDonnald-McGuinn for their critical review of the manuscript. Part of Figure 1 was created with BioRender.com.

## AUTHOR CONTRIBUTIONS

Conceptualization: MG, BH, JRV; Data curation: MG, BH, ES; Formal analysis: MG, BH; Funding acquisition: KDVB, JRV; Investigation: MG; Methodology: MG, BH; Project administration: KVDB, JRV; Resources: MG, JB, KDV, HP, GVB, HVE, KDVB; Software: BH, ES; Supervision: KVDB, JRV; Validation: MG, BH, ES; Visualization: MG, BH; Writing – original draft: MG, BH, KVDB, JRV; Writing – review and editing: MG, BH, ES, JB, KDV, HP, GVB, HVE, KVDB, JRV

## ETHICS AND CONSENT DECLARATION

This study was approved by the Ethical committee research UZ/KU Leuven (S65304 – S65991) and informed consent was obtained from all participants.

## CONFLIC OF INTEREST

The authors declare no conflicts of interest. MG presented her research at the ACLF conference 2023 for Oxford Nanopore Technologies. Travel costs were reimbursed by the company.

## ADDITIONAL FILES

**Supplementary table 1.**

**Patients’ phenotypes and variant classification**

Description of the phenotypes of the included patients, the underlying genomic (potential) pathogenic variants or VUS, and the classification of these variants according to the ACMG criteria [1], [2].

**Supplementary table 2.**

**DNA extraction and sequencing summary**

Description of the used methodologies for DNA extraction and sequencing as well as summary of the data output for each sample (estimated and basecalled bases as well as read length N50).

**Supplementary table 3**.

**Differentially methylated regions : quantification**

Mean methylation levels at six imprinted loci (first tab) and the *AR, RP2* and *PHF6* differentially methylated loci (second tab) for each haplotype of the six female patients’ blood samples (as well as the urine and buccal swab samples of EPI_13 for the loci on chromosome X). The values for which total coverage <10x or with <5x coverage for one of the haplotypes are represented in grey.

**Supplementary figure 1.**

**Differentially methylated regions : visualization**

Phased nanopore sequencing methylation levels at the same 6 imprinted loci (**Supplementary figure 1A**), as well as the *AR*, *RP2*, and *PHF6* genes (**Supplementary figure 1B**), in blood of the 6 female patients (as well as the urine and buccal swab samples of EPI_13 for the loci on chromosome X). Orange and blue represent the two different haplotypes of each sample. In the upper part of each Methylartist plot [3], the genome coordinates, genes and reads are illustrated. On each read, modified bases are represented by full, dark circles, unmodified bases by open circles. In the lower part of the plot, after translation from genome to CpG-only coordinate space, raw log-likelihood ratios are plotted above a smoothened graph of methylation fraction.

**Supplementary figure 2.**

**Nanopore methylation analysis allow to distinguish disease samples from controls and other diseases samples (t-SNE)**

t-SNE scatter plot from methylation levels at Sotos, Kabuki, Wiedemann-Steiner (WDSTS) and Cornelia de Lange (CdLS) syndromes episignature loci. Nanopore sequencing data of five healthy controls and seven patients with one of the four disorders are represented, along with the array-based reference [4] for the respective episignature. Patients sequenced with nanopore cluster in close proximity to their respective array-based episignature reference.

**Supplementary table 4. SVM classifiers results**

The excel sheets contains two tabs. The first one details the SVM Confidence Scores. In each row the values of the confidence score from each disorder’s SVM is shared for each sample. The second tab summarizes for each sample the highest confidence score (second row), and assigned disease (third row). Sample without recognized episignature (confidence score <0.3) are classified as control.

**Supplementary table 5**.

**Methylation values at known episignature loci : reference array data** [5] **and ONT data**

The excel sheet contains a tab for every evaluated disorder. For every disorder (tab), the first 2 columns indicate the chromosome and the end position of the CpG loci of the disorder specific episignature.

For every disorder specific episignature locus, Illumina derived methylation beta values in all disorders and controls shared in E. Aref-Eshghi *et al.* are shown (data was extracted from the provided supplementary table) [4]. The last 60 columns of every tab indicate the methylation values extracted from our Nanopore samples at every specific locus.

**Supplementary figure 3.**

**Phasing of methylome and genome variation**

Screenshots of the visualization of EPI_13’s Oxford Nanopore long read sequencing and Illumina short read sequencing BAM files in IGV [5] at two loci of interest: *PHF6* and *AR* genes. At the *PHF6* locus we observe the pathogenic c.306delA variant in cis with a maternally inherited SNV as well as methylation of the promotor region of the gene. At the *AR* locus we observe a paternally inherited SNV in *cis* with unmethylation of the promotor region of the gene.

