## Supplementary figures and images for "Nanopore sequencing-based episignature detection"

### Supplementary Figure 1a

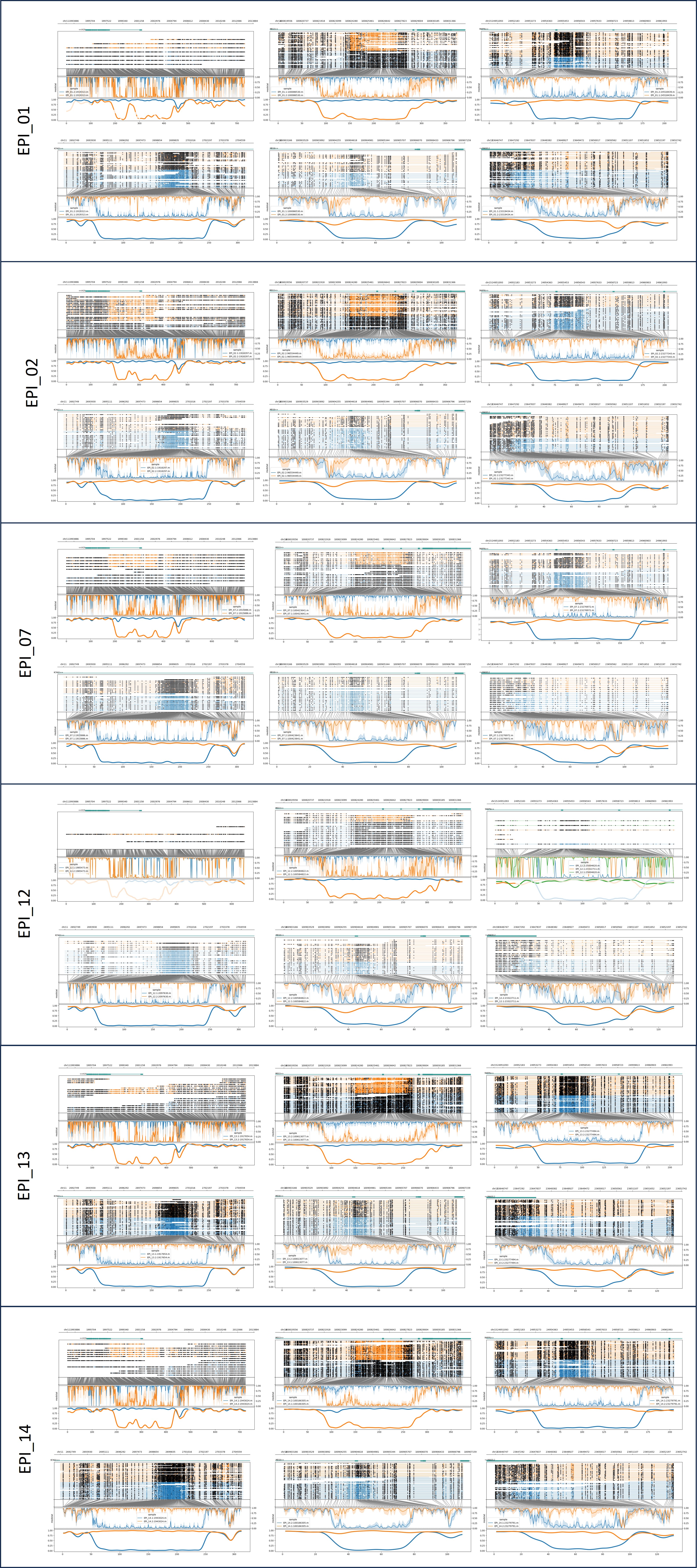

### Supplementary Figure 1b

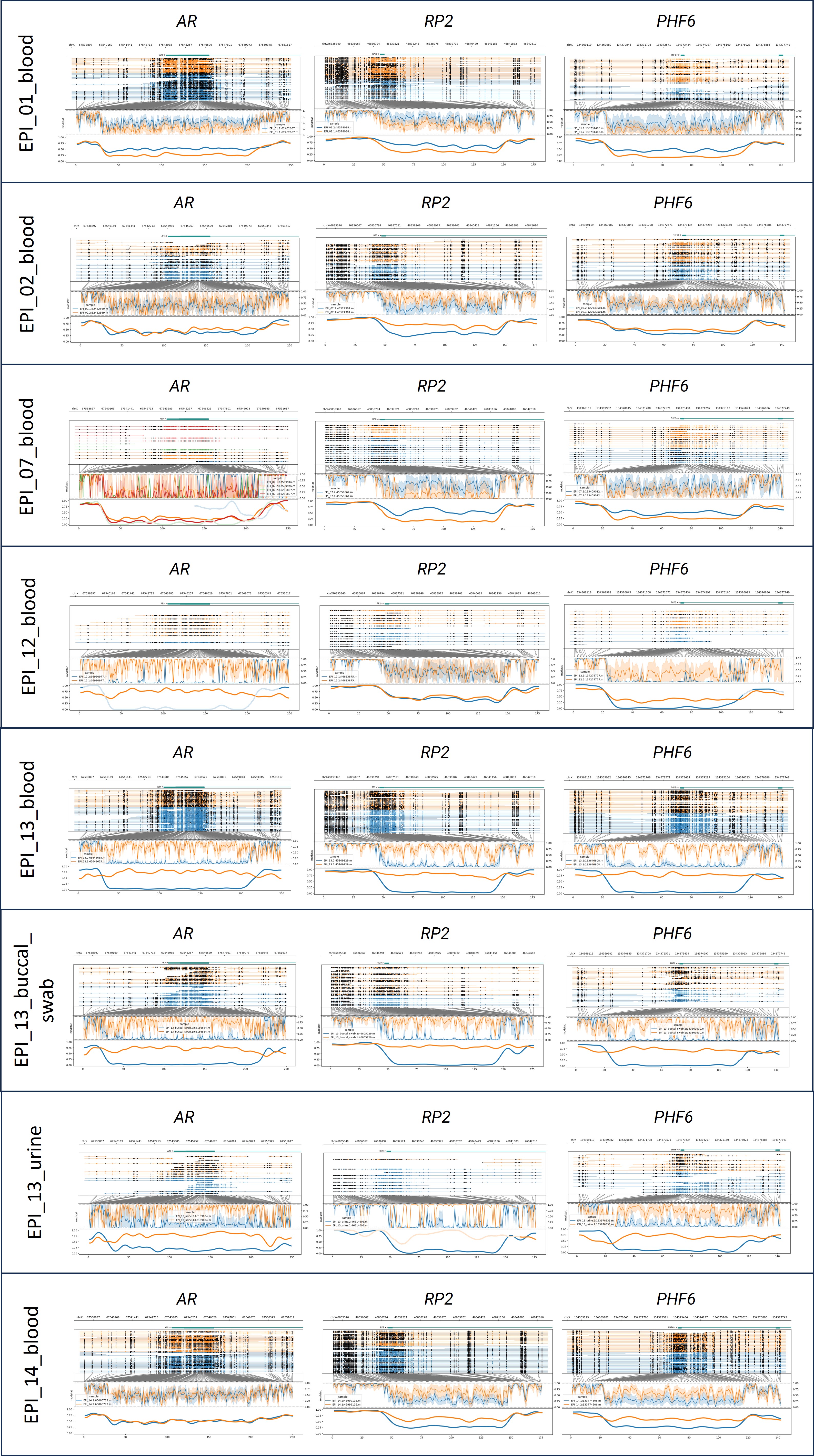

### Supplementary Figure 2

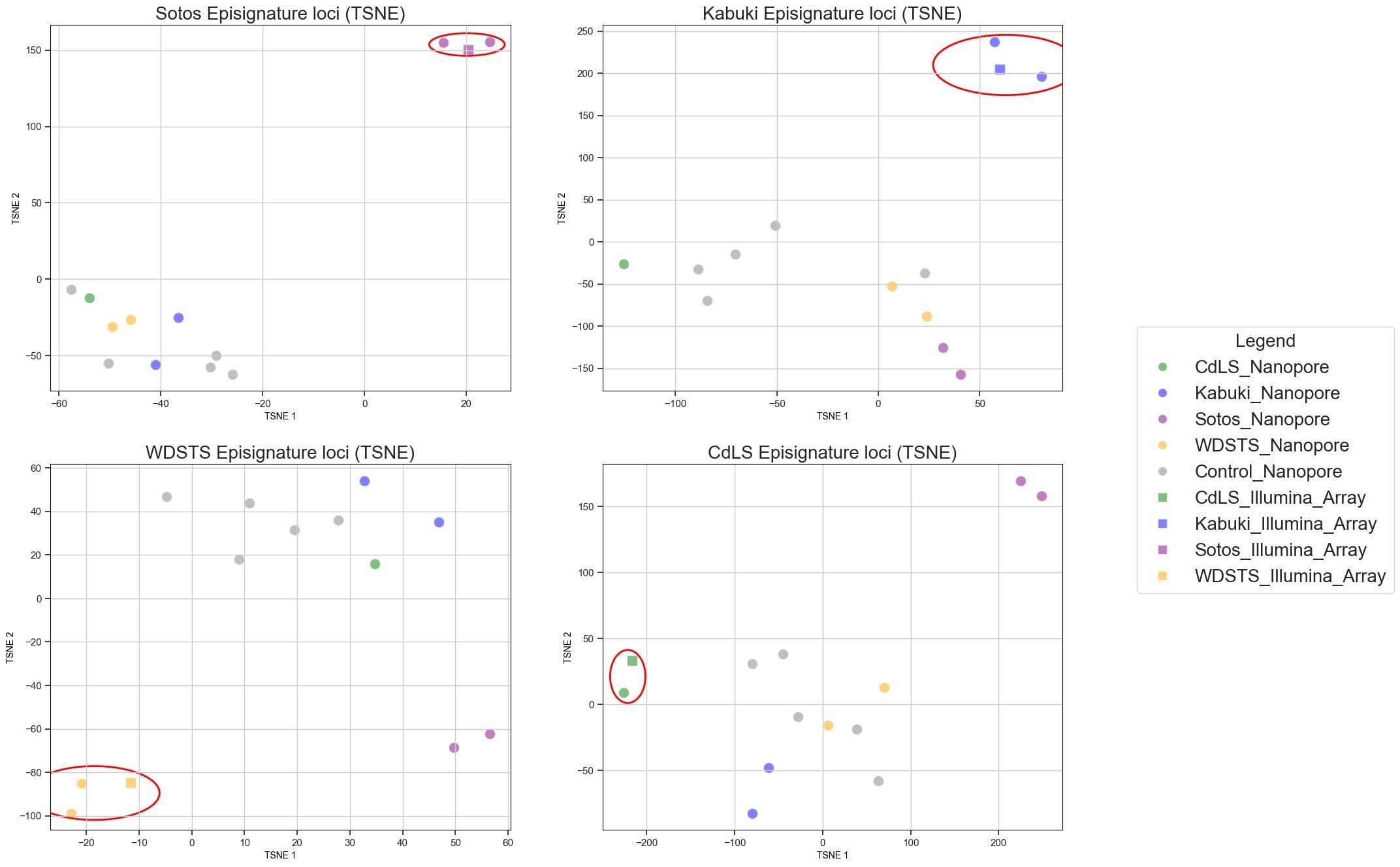

### Supplementary Figure 3

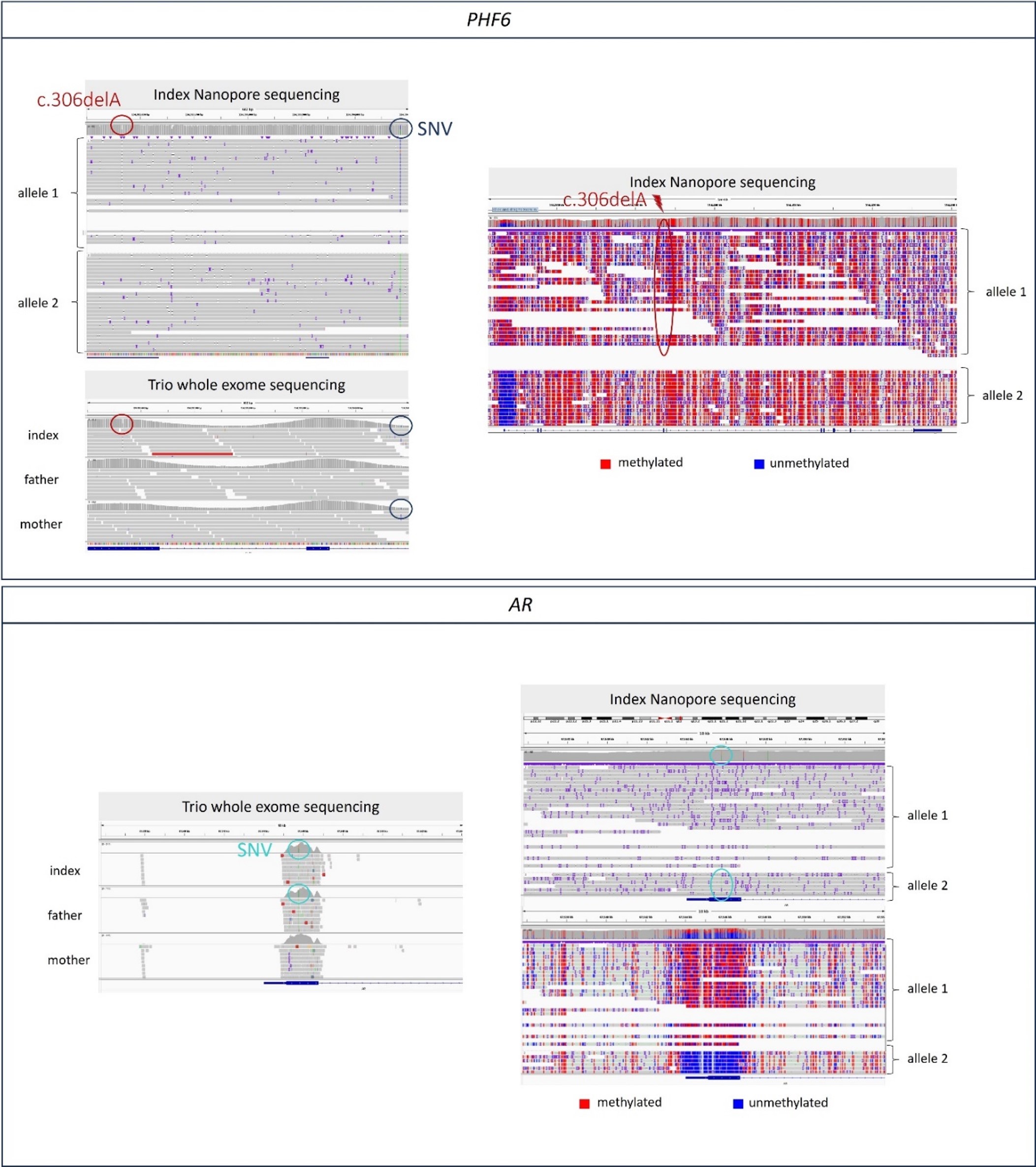
